# EpiSemoLLM: A Fine-tuned Large Language Model for Epileptogenic Zone Localization Based on Seizure Semiology with a Performance Comparable to Epileptologists

**DOI:** 10.1101/2024.05.26.24307955

**Authors:** Shihao Yang, Yaxi Luo, Neel Fotedar, Meng Jiao, Vikram R. Rao, Xinglong Ju, Shasha Wu, Xiaochen Xian, Hai Sun, Ioannis Karakis, Danilo Bernardo, Josh Laing, Patrick Kwan, Felix Rosenow, Feng Liu

**Author notes:** Senior Authors: TBD. **Corresponding Author:** Feng Liu, PhD, Department of Systems and Enterprises, Stevens Institute of Technology, 07030 Hoboken, United States.

## Abstract

**Significance:** Seizure semiology, the study of signs and clinical manifestations during seizure episodes, provides crucial information for inferring the location of epileptogenic zone (EZ). Given the descriptive nature of seizure semiology and recent advancements in large language models (LLMs), there is a potential to improve the localization accuracy of EZ by leveraging LLMs for interpreting the seizure semiology and mapping its descriptions to the corresponding EZs. This study introduces the *Epilepsy Semiology Large Language Model*, or *EpiSemoLLM*, the first fine-tuned LLM designed specifically for this purpose, built upon the Mistral-7B foundational model.

**Method:** A total of 865 cases, each containing seizure semiology descriptions paired with validated EZs via intracranial EEG recording and postoperative surgery outcome, were collected from 189 publications. These collected data cohort of seizure semiology descriptions and EZs, as the high-quality domain specific data, is used to fine-tune the foundational LLM to improve its ability to predict the most likely EZs. To evaluate the performance of the fine-tuned EpiSemoLLM, 100 well-defined cases were tested by comparing the responses from EpiSemoLLM with those from a panel of 5 epileptologists. The responses were graded using the rectified reliability score (rRS) and regional accuracy rate (RAR). Additionally, the performance of EpiSemoLLM was compared with its foundational model, Mistral-7B, and various versions of ChatGPT, Llama as other representative LLMs.

**Result:** In the comparison with a panel of epileptologists, EpiSemoLLM achieved the following score for regional accuracy rates (RAR) with zero-shot prompts: 60.71% for the frontal lobe, 83.33% for the temporal lobe, 63.16% for the occipital lobe, 45.83% for the parietal lobe, 33.33% for the insular cortex, and 28.57% for the cingulate cortex; and mean rectified reliability score (rRS) 0.291. In comparison, the epileptologists’ averaged RAR scores were 64.83% for the frontal lobe, 52.22% for the temporal lobe, 60.00% for the occipital lobe, 42.50% for the parietal lobe, 46.00% for the insular cortex, and 8.57% for the cingulate cortex; and rectified reliability score (rRS) with mean of 0.148. Notably, the fine-tuned EpiSemoLLM outperformed its foundational LLM, Mistral-7B-instruct, and various versions of ChatGPT and Llama, particularly in localizing EZs in the insular and cingulate cortex. EpiSemoLLM offers valuable information for presurgical evaluations by identifying the most likely EZ location based on seizure semiology.

**Conclusion:** EpiSemoLLM demonstrates comparable performance to epileptologists in inferring EZs from patients’ seizure semiology, highlighting its value in epilepsy presurgical assessment. EpiSemoLLM outperformed epileptologists in interpreting seizure semiology with EZs originating from the temporal and parietal lobes, as well as the insular cortex. Conversely, epileptologists outperformed EpiSemoLLM regarding EZ localizations in the frontal and occipital lobes and the cingulate cortex. The model’s superior performance compared to the foundational model underscores the effectiveness of fine-tuning LLMs with high-quality, domain-specific samples.

## 1 Introduction

Epilepsy is one of the most common neurological diseases, affecting more than 70 million people worldwide [1]. Each year, approximately 50.4 per 100,000 people develop new-onset epilepsy [2]. At present, the primary treatment for epilepsy is antiseizure medications, which successfully controls seizures in about two-thirds of patients [3]. However, for those with drug-resistant focal epilepsy, surgical resection/disconnection of the epileptogenic zone (EZ) may provide a curative solution. *Seizure semiology* - the study of signs and symptoms exhibited and experienced by a patient during epileptic seizures [4] - offers valuable clues for the inference of Symptomatogenic Zone (SZ) [5] thus indicative of the EZs [6]. Interpretation on the initial ictal symptom and the evolution of ictal semiology can help in inferring the localization of the EZ, which can serve as a crucial step for preoperative assessment and help to achieve optimal surgical outcomes.

Given the limited surgical epilepsy cases at each epilepsy center, it is hard to summarize and generalize the systematic knowledge on mapping from the descriptions of seizure semiology to EZs, complicated by (1) unstructured and subjective textual descriptions of seizure semiology by different epileptologists; (2) conflicting lateralizing or localizing signs occurring in a single seizure [6]; (3) the bias on presumption of the temporal lobe and frontal lobe epilepsy. This problem can be potentially addressed by aggregating a large cohort of data with descriptions of seizure semiology paired with the validated EZs and a generalizable machine learning model to extract knowledge on seizure semiology interpretation to mitigate the inconsistencies caused by different epileptologists and different epilepsy centers.

Recently, large language models (LLMs) have demonstrated their capabilities across a wide range of natural language processing (NLP) tasks [7]. ChatGPT, a representative example, is trained on a diverse database of text from various domains [8] and can emulate human experts with cross-disciplinary knowledge. In medical informatics, ChatGPT exhibits an advanced proficiency in processing and interpreting extensive textual data, making it a valuable tool for information retrieval, clinical decision support, and medical report generation [7, 9, 10, 11]. However, previous studies have raised concerns about the responses from LLM trained from the general public data, especially its poor performance in highly specialized domains due to limited data [12, 13]. To address this, fine-tuning LLM on locally collected and well-annotated data is a common approach that requires relatively low computational resources. Examples include ChatDoctor, which is a fine-tuned LLM based on Llama and trained on a dataset of 100,000 patient-doctor dialogues, providing informative, accurate, and professional advice [14]. Xie et al. created Me-LlaMA by fine-tuning Llama using an instruct-tuning medical dataset, achieving superior performance in both general and medical tasks [15]. Labrack et al. introduced the BioMistral, fine-tuned on medical data from PubMed Central, which demonstrated the state-of-the-art (SOTA) performance in medical question-answering tasks [16].

The growing applications of LLM enriched by domain specific data records in a variety of clinical applications inspired this study to develop a fine-tuned LLM on interpreting seizure semiology by mapping its descriptions to the underlying EZs, which can serve as a valuable decision making tool during presurgical workup Phase I to (1) provide accurate localization of EZs based on seizure semiology, (2) streamline the decision making process and shorten the time to definitive treatment, (3) help reduce mistakes and treatment bias especially in the resource-limited epilepsy centers, (4) reduce the inter-rater variability on the interpretation of seizure semiology.

In this study, we used a collected dataset of seizure semiology paired with validated EZs to fine-tune an LLM to improve its EZ localization performance. Mistral-7B, an emerging lightweight LLM, has outperformed larger models like Llama-2-13B in various aspects and is suitable for fine-tuning domain-specific tasks [17]. We selected the Mistral-7B-Instruct-v0.3 model as the foundation model for the EZ localization, resulting in the fine-tuned model termed as EpiSemoLLM. The framework of EpiSemoLLM is illustrated in **Figure 1**. To evaluate EpiSemoLLM’s advantages and limitations, we systematically compare its performance with human experts and other SOTA LLMs, such as different versions of ChatGPT.

**Figure 1:**
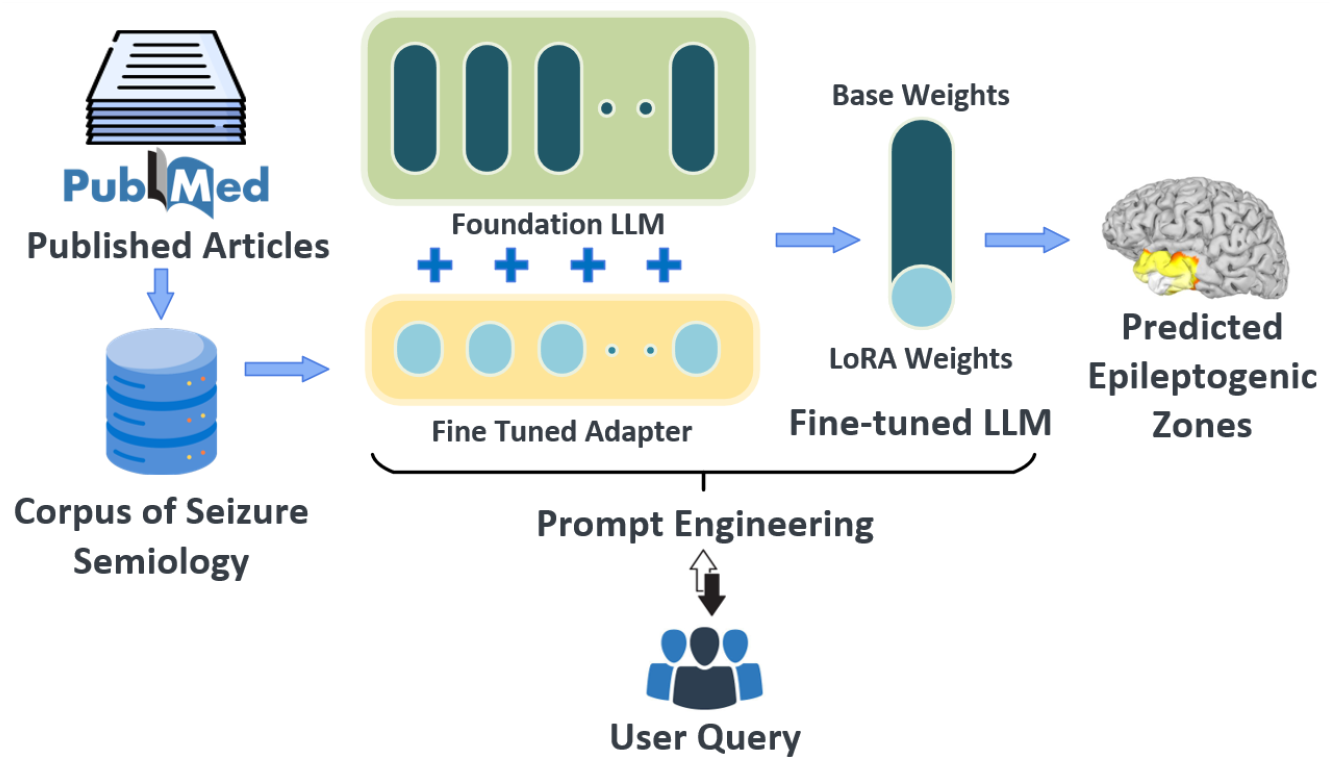
Framework for fine-tuning LLMs to predict EZs based on seizure semiology.

## 2 Methods

### 2.1 Seizure semiology and EZ data collection and annotation

We compiled an epilepsy-specific database from published articles indexed by PubMed. By searching for terms such as “seizures”, “epilepsy”, “clinical semiology”, and “seizure semiology”, we identified 189 articles [18]. These selected articles documented over 900 epilepsy cases, detailing seizure semiology across various surgically validated EZs. The confidence in the epileptogenic zone location reported locations (i.e., ground truth) was labeled based on postoperative outcomes (defined as seizure freedom after surgery), concordance of imaging and neurophysiology, or available stereoelectroencephalography (sEEG) findings, as recently suggested by Ryvlin *et al*. [31]. Cases with low levels in confidence of EZ localization or nonspecific seizure semiology were excluded. For example, cases indicating only hemisphere-level EZs, like right hemispherectomy or left subtotal hemispherectomy, without lobe details or specific regional details, were excluded. Additionally, cases described with nonspecific terms, like ‘non-specific aura’ or those aggregating a large patient cohort without providing detailed semiology for each individual, were excluded.

A thorough collection of patient data was compiled, encompassing demographics, seizure semiology, imaging and/or sEEG findings (if provided), and surgical results. The resultant database comprises 865 patient-derived semiology-EZ pairs, with a demographic spread of 134 right-handed, 22 left-handed, 3 ambidextrous, and 706 unspecified-handed individuals, aged from newborn to 77 years. These semiology-EZ samples include both single and multiple EZ locations. A PRISMA flow chart for dataset construction and the finalized regional distribution of screened records are listed in **Fig. 2** and **Fig. 3**, respectively. In documenting these locations, we have referenced a classification system based on FreeSurferWiki and LCN-CortLobes [20]. This system organizes the EZs into six broad regions: frontal lobe, parietal lobe, temporal lobe, occipital lobe, cingulate cortex, and insular cortex [21].

**Figure 2:**
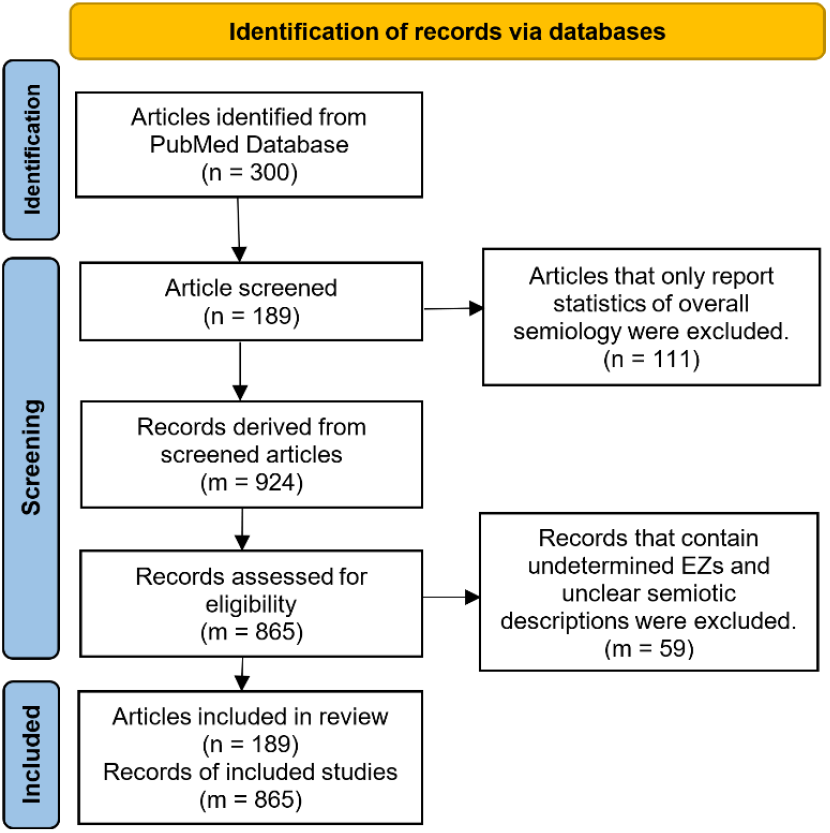
PRISMA flow chart for dataset construction

**Figure 3:**
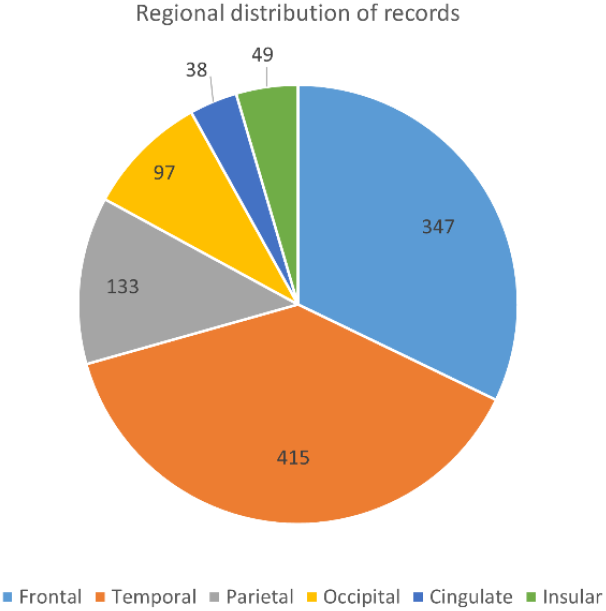
Regional distribution of screened records.

### 2.2 Fine-tuning of LLM

In this study, the collected seizure semiology records from our self-compiled database were leveraged to investigate the capabilities of a fine-tuned LLM in identifying the most likely locations of the EZs. We used 765 pairs of semiology descriptions and validated EZs for the fine-tuning of a light-weighted foundational backbone model, which is the Mistral-7B, due to its good performance at a variety of tasks [17]. The remaining 100 records were used as the testing dataset to evaluate EpiSemoLLM, ChatGPT, and a panel of epileptologists.

The model was fine-tuned using a self-instructed format dataset similar to Stanford Alpaca [22]. To minimize the impact of fine-tuning on the overall LLM performance and improve the fine-tuning efficiency, we applied Low-Rank Adaption (LoRA) [23], integrated into the Hugging Face Parameter Efficient Fine-Tuning (PEFT) library. Instead of comprehensively fine-tuning the entire weight matrix of the pre-trained LLM, LoRA fine-tunes two smaller matrices that approximate the larger weight matrix. These matrices construct the LoRA adapter, which, after fine-tuning, can be easily loaded into pre-trained models and used for inference.

The key hyperparameters used for fine-tuning include the epochs number being set to be 2, a learning rate of 3 × 10^−5^, a maximum sequence length of 512 tokens, a weight decay of 0.001, a warmup ratio of 0.03, a LoRA alpha to be 32, and a LoRA rank of 16. The prompt format can be classified in two ways: zero-shot prompting (ZSP) fine-tuning [24], where the model was asked to identify EZs solely based on semiology reports, and few-shot prompting (FSP) [25], which differed from ZSP by providing three sample hints as prior knowledge to assist model training.

### 2.3 Semiology Interpretation Data Collection

To compare the performance of LLMs (both foundation models and fine-tuned ones) and epileptologists on the interpretation of seizure semiology, we invited a panel of eight epileptologists, with an average of 10 years of experience in treating patients with epilepsy, to complete an online survey on seizure semiology interpretation with unconstrained time and attempts (https://survey.zohopublic.com/zs/NECl0I). In this survey, we used the 100 hold-out semiology records, spanning all six regions from our self-compiled database, to gather the epileptologists’ opinions on the most likely EZs. The selection of semiology records met the following criteria, reviewed by the epileptologists: (1) the records provided comprehensive and explicit descriptions of seizure signs and symptoms; (2) the distribution of EZs covered all six general regions, rather than being focused on a particular region; and (3) the records captured the broadest possible range of seizure symptoms.

All survey responses were collected from January 2024 to February 2024. During this period, 70 survey invitations were sent to doctors specializing in epilepsy, identified from the National Association of Epilepsy Center, American Epilepsy Society, and International League Against Epilepsy (ILAE). Among these, five epileptologists completed the survey in full, and three others partially completed it. This can also highlight the advantage of all-time availability and non-fatigue features of the clinical AI models.

In addition to comparing the performance of the fine-tuned LLM to epileptologists, we also evaluated the performance of non-fine-tuned pre-trained LLMs, including Mistral-7B-instruct, ChatGPT-3.5, and ChatGP-4.0, as the baseline models. For ChatGPT prediction, we adopted a stringent methodology where each query was entered in a separate “New Chat” session to mitigate any potential bias or interference due to the stateful nature of ChatGPT.

### 2.4 Statistical Analysis

All responses from EpiSemoLLM, foundation LLMs (ChatGPT-3.5 and -4.0, Mistral-7B-instruct-v0.3, Llama-2-7B-chat-hf, Llama-2-13B-chat-hf, Llama-3-8B-Instruct), and the panel of epileptologists underwent thorough review, with the addressed EZ locations being systematically cataloged and summarized. By evaluating and comparing responses from EpiSemoLLM, baseline foundation LLMs, and epileptologists, an in-depth understanding of the strengths and limitations of AI-generated medical information can be achieved, which provides insights into its future applications in the biomedical and healthcare domains.

To provide a quantitative assessment of the responses provided by the LLMs and epileptologists, two statistical performance metrics were introduced: the rectified Reliability Score (rRS) and the Regional Accuracy Rate (RAR) [26].

The introduced rRS quantifies the accuracy of responses, where a score of 1 indicates 100% correct identification of EZ locations, aligning perfectly with the ground truth. A score between 0 and 1 suggests partially accurate, while a score less than 0 indicates misleading responses, potentially complicating preoperative evaluations by neurologists. The rRS is calculated as follows:

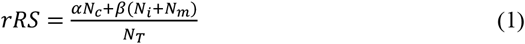

where *N*_*T*_ denotes the total count of different EZs identified in both the ground truth and the model predictions for a given semiology case. *N*_*c*_ and *N*_*i*_ respectively represent the counts of correct and incorrect predictions made by LLMs or epileptologists, given the ground truth of each epilepsy case, while *N*_*m*_ is the number of EZs in the ground truth that was not identified by either LLM or epileptologists. Correct predictions (*N*_*c*_) positively contribute with a weight of *α*=1, while incorrect ones (*N*_*i*_) and missed ones (*N*_*m*_) carry a negative weight of *β*=−0.5 to account for their potential to mislead epileptologists in determining the correct EZ location.

The weighting factors (*α*=1, *β*=−0.5) are chosen to balance the impact of correct and incorrect responses. With *α*=1 and *β*=0, the rRS would be greater than or equal to zero, which simply represents the proportion of correct answers but could not reflect the potentially negative impact of incorrect and misleading answers. Conversely, with *α*=1 and *β*=−1, equal weight is assigned to both positive and negative impacts. However, this would result in an rRS of zero when the count of correct responses matches those of incorrect ones for a given semiology, failing to convey the positive impact of correct identifications in narrowing down the diagnoses of EZ. In our study, we set *α*=1, *β*=−0.5 to prevent the complete negation of a correct answer by an incorrect one, thus ensuring that the rRS accounts for the influence of both positive and negative responses on clinical decision-making, and with these weighting factors, the range of rRS is [-0.5, 1.0].

For example, if we consider a semiology case where the ground truth for the EZ is the frontal, parietal, and occipital lobes, but LLM’s response identifies the frontal, temporal, and parietal lobes as the potential EZs. In this scenario, two correct identifications (*N*_*c*_=2), one incorrect identification (*N*_*i*_=1), and one missing prediction (*N*_*m*_=1), with the ground truth and identification specifying four different EZ (*N*_*T*_=4), result in an rRS of 0.25.

The RAR is introduced as a region-specific metric designed to evaluate the precision of EZ localization for specific regions. It calculates the region-level localization performance by LLMs or epileptologists against the ground truth. The RAR values range from 0% to 100%. When the response from the LLMs or epileptologist perfectly matches the ground truth, the RAR is 100%; otherwise, it is less than 100%.

For a region *x*, which is one of the following regions: frontal lobe, parietal lobe, temporal lobe, occipital lobe, cingulate cortex, and insular cortex, the RAR(*x*) is calculated as:

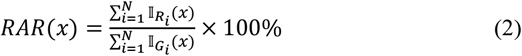

with

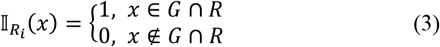

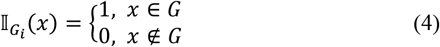

where *N* represents the number of semiology-EZ pairs; *G* and *R* represent the sets of EZ locations from the ground truth and the responses from LLM or epileptologists, respectively. 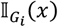 is an indicator function that scores 1 if the general region *x* is included in the ground truth *G* of a semiology-EZ pairing; otherwise, it is 0. 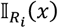 is a similar indicator function that scores 1 if region *x* is correctly identified in the intersection of the response *R* and the ground truth *G* for a semiology-EZ pairing and 0 otherwise. 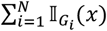 quantifies the aggregate presence of the general region *x* within the ground truth in *N* semiology-EZ pairs. 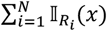 quantifies the intersection of the response *R* and the ground truth *G* for a semiology-EZ pairing in *N* semiology-EZ pairs, regardless of whether a single region or multiple regions for each pair.

For instance, we can calculate the RAR for the frontal lobe within a dataset of 100 semiology-EZ pairs. In this dataset, the ground truth may include both cases with a single EZ located in the frontal lobe and cases where the frontal lobe is one of multiple EZs. Suppose there are 50 instances where the ground truth indicates the frontal lobe as an EZ; the value of 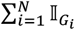 (*Frontal Lobe*) would be 50. If only 30 responses — whether from LLM or an epileptologist — correctly match the ground truth, the value of 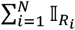 (*Frontal Lobe*) would be 30. Consequently, the RAR (Frontal Lobe) would be calculated as 60%.

## 3 Results

### 3.1 Comparison of Responses from EpiSemoLLM and Multiple LLMs

In this study, the EpiSemoLLM was fine-tuned based on 765 records and evaluated on a hold-out set of 100 records, as detailed in 2.2. Pre-trained LLMs (LLM without fine-tuning), specifically Mistral-7B, ChatGPT-3.5, and ChatGPT-4.0 were used as baseline models for comparison.

In EZ location inference using ZSP, EpiSemoLLM demonstrated the highest rRS score with a mean of 0.291 compared to the average and the best epileptologist’s performance with mean rRS of 0.148 and 0.192, respectively, and the best baseline result from ChatGPT-4.0 with mean rRS of 0.195.

For the six regions, the RARs for EpiSemoLLM with ZSP were: 60.71% for the frontal lobe, 83.33% for the temporal lobe, 63.16% for the occipital lobe, 45.83% for the parietal lobe, 33.33% for the insular cortex, and 28.57% for the cingulate cortex. With FSP, the RARs for EpiSemoLLM were: 39.29% for the frontal lobe, 75.00% for the temporal lobe, 73.68% for the occipital lobe, 62.50% for the parietal lobe, 33.33% for the insular cortex, and 14.29% for the cingulate cortex.

By contrast, Mistral-7B with ZSP had RARs of: 57.14% for the frontal lobe, 80.56% for the temporal lobe, 15.79% for the occipital lobe, 12.50% for the parietal lobe, 0.00% for the insular cortex, and 0.00% for the cingulate cortex. With FSP, Mistral-7B had RARs of: 21.43% for the frontal lobe, 69.44% for the temporal lobe, 57.89% for the occipital lobe, 37.50% for the parietal lobe, 0.00% for the insular cortex, and 0.00% for the cingulate cortex. These results highlight the importance of fine-tuning with highly domain-specific datasets.

Besides, the best result from that ChatGPT family was ChatGPT-4.0 with ZSP, which had RARs of: 79.31% for the frontal lobe, 69.44% for the temporal lobe, 57.89% for the occipital lobe, 54.17% for the parietal lobe, 0.00% for the insular cortex, and 0.00% for the cingulate cortex. With FSP, the RARs were: 79.31% for the frontal lobe, 63.89% for the temporal lobe, 63.16% for the occipital lobe, 45.83% for the parietal lobe, 0.00% for the insular cortex, 14.29% for the cingulate cortex.

These results show that EpiSemoLLM performs well in interpreting seizure semiology not only for the common frontal and temporal lobe epilepsies but also for the less common occipital and parietal cases. Even in rare areas, i.e., cingulate and insular cortex, according to the regional distribution from the screened records shown in **Fig. 3**, EpiSemoLLM provides more accurate inferences than baseline models.

### 3.2 Comparison of Responses from EpiSemoLLM and Epileptologists

An online survey comprising 100 questions regarding EZ locations and corresponding seizure semiology was conducted to gather responses from eight Board-certified epileptologists. Out of these, five participants completed the survey entirely, while three others completed it partially. Consequently, the analysis focused on the fully completed responses from five epileptologists (E1, E2, E3, E4, E5). After excluding ambiguous characterization of EZs from the literature, the analysis was further narrowed down to 90 questions for the comparison between all LLMs and epileptologists.

In the general region localization, epileptologist E5 demonstrated the highest rRS with a mean of 0.192 (**Fig. 4**). For six regions, the mean RARs were: 64.83% for the frontal lobe, 52.22% for the temporal lobe, 60.00% for the occipital lobe, 42.50% for the parietal lobe, 46.00% for the insular cortex, and 8.57% for the cingulate cortex, while the best possible RAR from all epileptologists’ responses was 82.76% for the frontal lobe, 50.00% for the temporal lobe, 68.42% for the occipital lobe, 41.67% for the parietal lobe, 40.00% for the insular cortex, and 0.00% for the cingulate cortex (**Fig. 5**).

**Figure 4:**
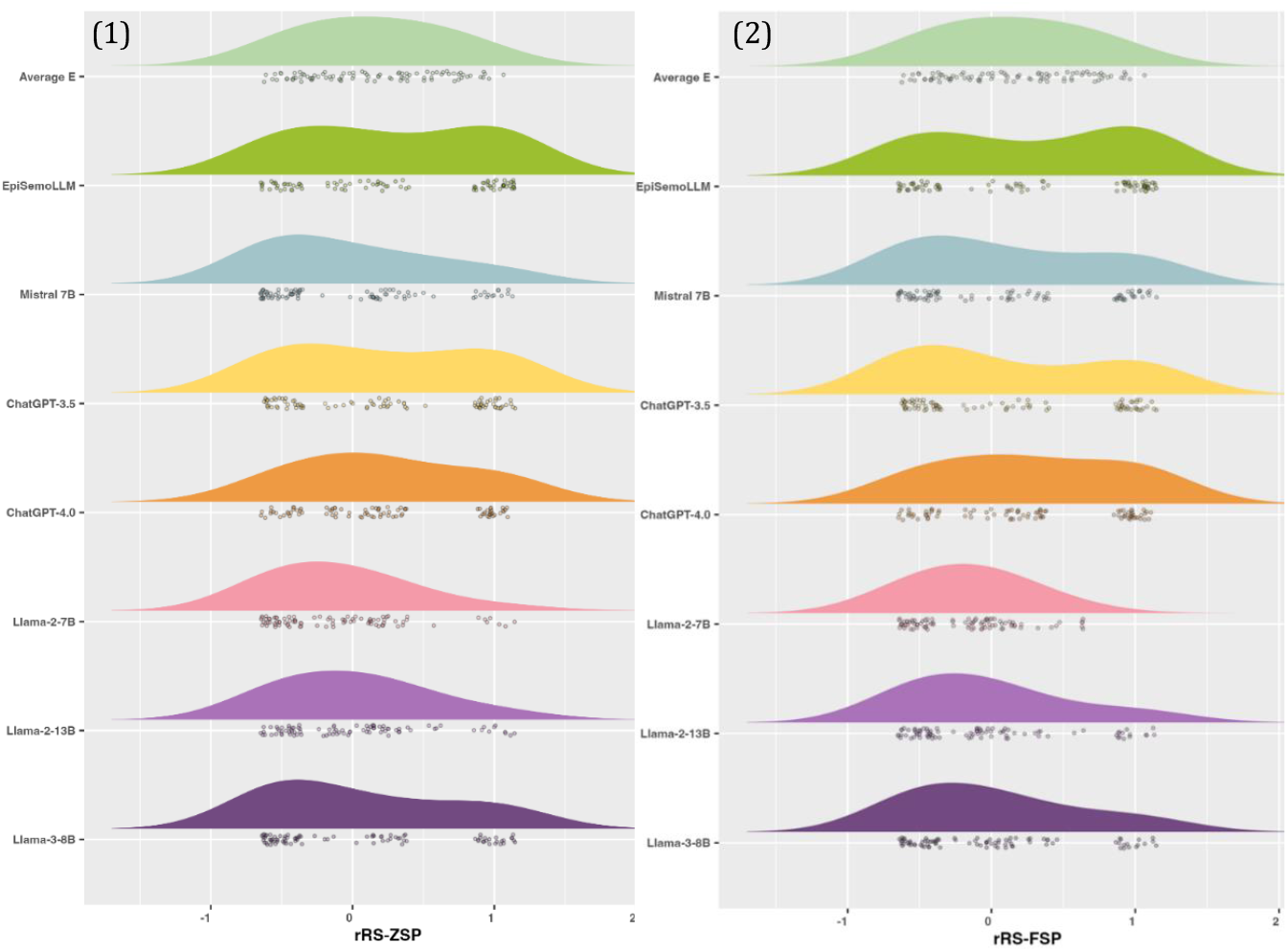
(1) Distribution of average performance of epileptologists and other SOTA LLMs with zero-shot prompting according to rRS metrics. (2) Distribution of average performance of epileptologists and other SOTA LLMs with few-shot prompting performance according to rRS metrics.

**Figure 5:**
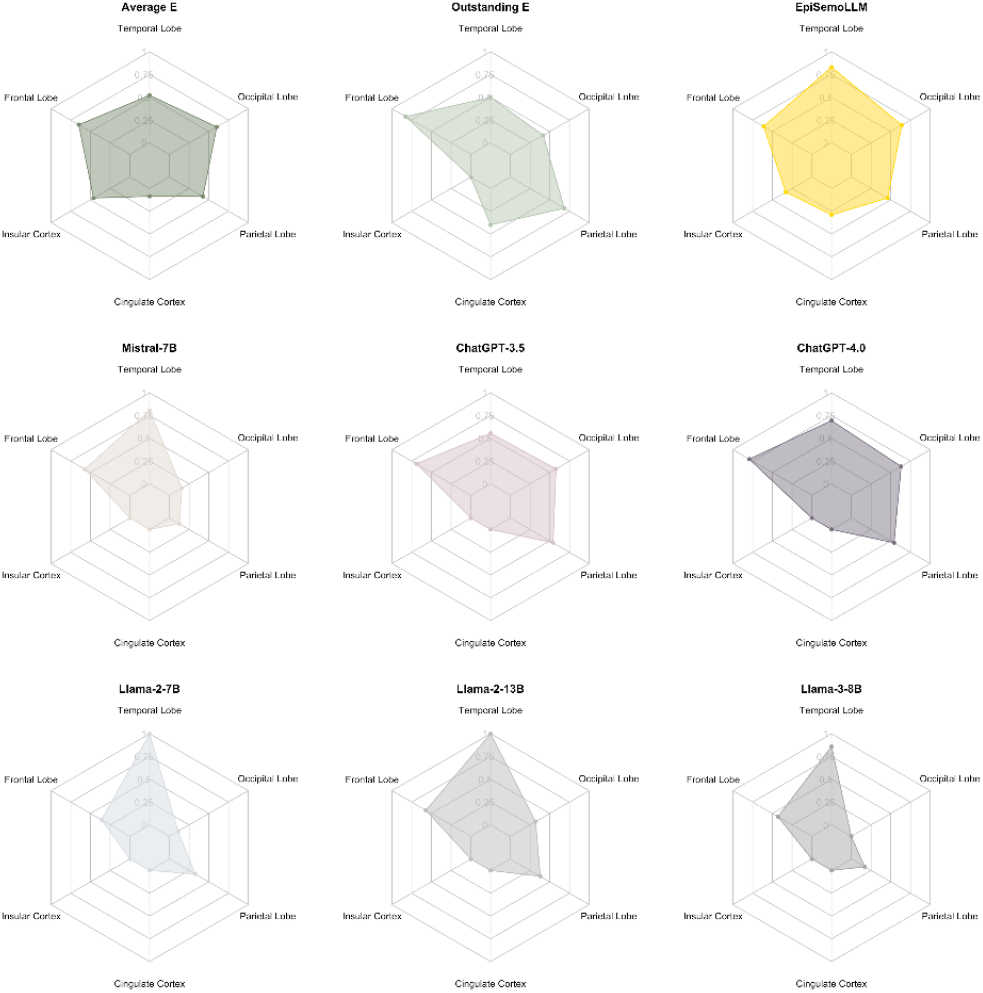
RAR scores for average performance of epileptologists and other SOTA LLMs with zero-shot prompting.

Notably, EpiSemoLLM demonstrated comparable or superior accuracy to the best possible answers from all epileptologists in interpreting seizure semiology related to the frontal, temporal, parietal, and occipital lobes, as well as rare regions, the insular and cingulate cortex. (**Fig. 5)**. The FSP results are presented in supplementary material.

A group *t*-test based on the 100 times bootstraps was applied to identify the significance of the difference in RAR performance between the averaged epileptologists and EpiSemoLLM with different types of prompting, as shown in **Fig. 7**. The results indicate that EpiSemoLLM is comparable to or even outperforms epileptologists in frontal, temporal, parietal, occipital, and cingulate regions.

**Figure 7:**
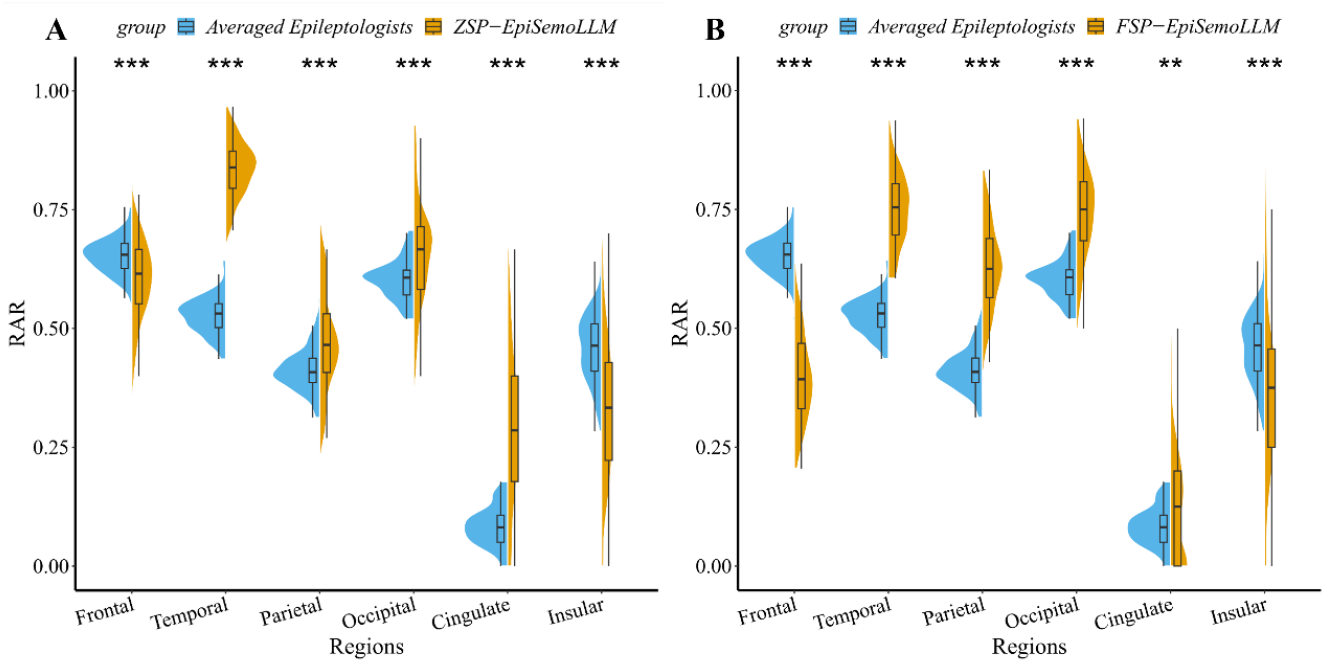
T-test significance of RAR between epileptologists and EpiSemoLLM with zero-shot prompting (A) and few-shot prompting (B) on different regions, based on the 100 times bootstraps. Stars indicate significance levels: - (no significance), * (p-value<0.05), ** (p-value<0.01), and *** (p-value<0.001).

However, in the insular cortex identification, the EpiSemoLLM performs poorer than epileptologists, while the gap is not too large. A visualization of RAR distribution across different brain general regions is provided in **Fig 8**.

**Figure 8:**
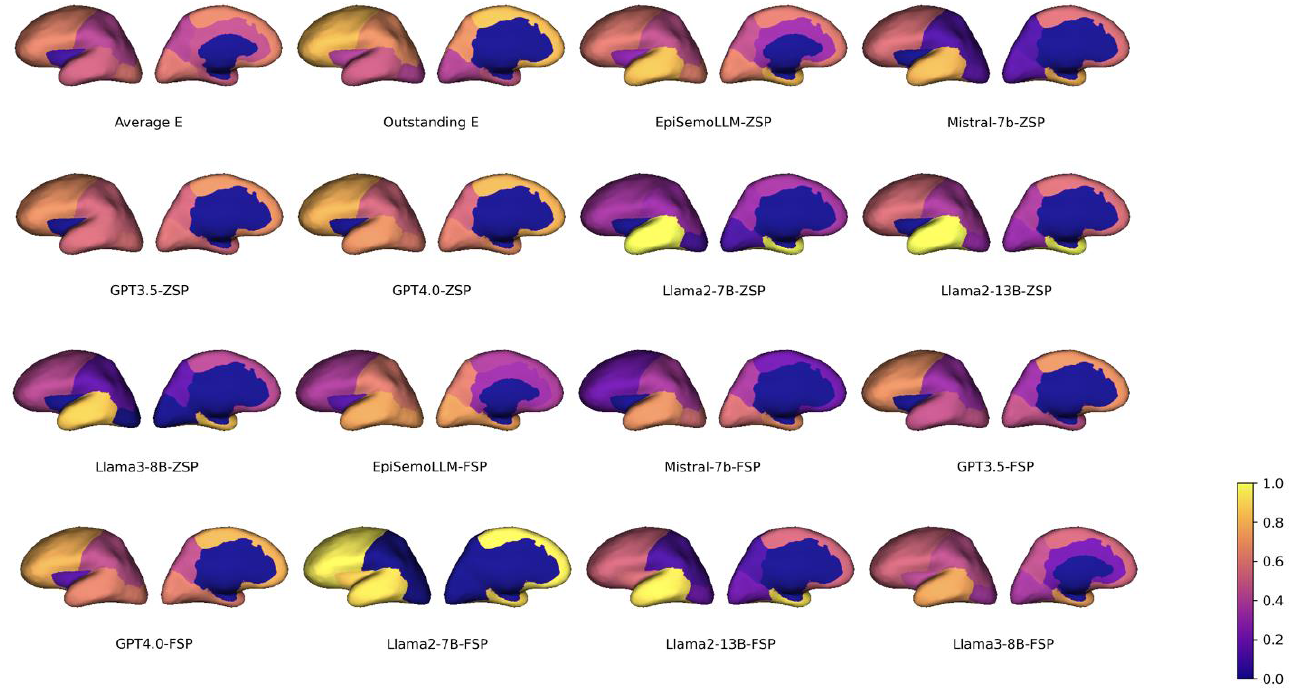
RAR distribution across different brain regions.

## 4 Discussion

This study proposes the first fine-tuned LLM for predicting EZ locations by interpreting seizure semiology. The fine-tuned model, EpiSemoLLM, was systematically compared with a cohort of board-certified epileptologists and a few foundational LLMs, including ChatGPT, Llama and Mistral. Trained from a meticulously collected cohort of seizure semiology-EZs pairs from published literature, EpiSemoLLM showed improved performance over the original foundation model, highlighting the importance of fining-tuning using carefully annotated and domain specific data.

EpiSemoLLM was evaluated using metrics of rRS and RAR, which was constructed to balance the score of correct answer while penalizing the score with misleading answers. The rRS evaluation revealed that EpiSemoLLM demonstrated a performance comparable to a panel of epileptologists, who provided responses to all 100 cases overcoming limitations in time and motivation, typical restrictions to human performance. Moreover, it outperformed its original version, Mistral-7B-instruct, underscoring the importance of fine-tuning with domain-specific datasets. Results also indicated that ZSP provided more robust and accurate results compared to FSP. Few-shot prompting can introduce biases when training samples are insufficient, leading to less accuracy for samples similar to the previous given cases.

Compared to the heavyweight ChatGPT models, there is a significant gap in both knowledge reserve from pre-training and model inference capabilities. However, according to the RAR assessment, EpiSemoLLM exceeds all baseline models in inference accuracy across most epileptogenic regions and showed significant improvements in some uncommon regions, particularly the insular cortex. Compared to the results from epileptologists, EpiSemoLLM achieved performance comparable to or better than the average level of epileptologists, though there remains a gap in performance for the cingulate cortex. This discrepancy is attributed to the limited number of training samples (only 29) for the cingulate, compounded by the overlap in semiology with other regions. Even among experts, the localization accuracy for the cingulate cortex is the lowest, with an average RAR of less than 10%.

Our results align with findings from previous studies assessing LLM performance in epilepsy-related inquiries. Specifically, Kim et al. evaluated the reliability of responses of LLM to 57 commonly asked questions about epilepsy symptoms and diagnosis, with all responses reviewed by two epileptologists. The results indicated that almost all questions were either of “sufficient educational value” response or “correct but inadequate” response [27]. Wu et al. assessed LLM performance on 378 epilepsy-related questions and 5 questions related to emotional support, finding that LLM provided “correct and comprehensive” answers to 68.4% of the questions but performed poorly on “prognostic questions”, with only 46.8% rated as comprehensive [28].

The improvement in LLM performance with fine-tuning is consistent with previous work. Li et al. fine-tuned Llama with patient-doctor dialogues, significantly enhancing its ability to provide reliable medical information [14]. Wu et al. performed instruct tuning on Llama for medical question-answering, with the fine-tuned LLM exhibiting superior performance, even surpassing more complex state-of-the-art models such as ChatGPT [29].

While this study offers valuable insights into EpiSemoLLM’s capability to provide reliable interpretations of seizure semiology, it has several limitations. First, the training sample size is limited for some specific regions and is unbalanced among regions. Additionally, different EZ locations may share similar semiology, complicating the fine-tuning process. Unlike ChatGPT, which tends to provide relatively conservative answers, EpiSemoLLM boldly gives the most likely inference, even if it may be incorrect, based on its fine-tuning samples. The overlap of symptoms in different EZs allows EpiSemoLLM to learn a broader range of possibilities, resulting in it offering more potential EZs based on the semiology descriptions. The collected data can only allow us to infer the lobe level EZs, given the most of the specific brain regions are missing. However, with more data available in the future, we believe more specifical region localization is feasible. Finally, it is seizure semiology that is more directly linked to the SZ which is indicative of EZ, which limits the capability to localize the EZ with 100% accuracy. Nevertheless, the EpiSemoLLM can provide valuable information during Phase 1 of presurgical workup. In order to allow optimal EZ location prediction a LLM would have to also include data from imaging (MRI and PET), interictal and ictal EEG, neuropsychology etc., as generally needed in presurgical epilepsy diagnosis (Rosenow and Lüders 2001).

However, it is crucial to recognize that the information provided by EpiSemoLLM may not always be supported by reliable sources, posing a challenge to verifying its responses. Furthermore, medical professionals, including epileptologists and neurosurgeons, must fully recognize the limitations of EpiSemoLLM and exercise caution when utilizing its responses. Future work should aim to address these issues by improving the training sample size, diversity, and quality. Additionally, more advanced LLM architectures with larger weights can be applied to achieve better overall performance after fine-tuning [30]. Finally, the current version of EpiSemoLLM is based solely on textual information on semiology. Thus, future studies could explore the feasibility of using both semiology descriptions, video and neuroimaging data from other modalities for EZ localization, offering a novel method for preoperative assessments.

## 5 Conclusion

In this cross-sectional study of seizure semiology interpretation, EpiSemoLLM, the first fine-tuned LLM specializing on prediction of EZs based on the description of seizure semiology, demonstrated performance comparable to epileptologists and improved performance than other foundational LLMs. The model excelled in regions where EZs are commonly located and showed significant improvement in insular EZ locations compared to other SOTA LLMs. Our results demonstrate the feasibility of utilizing LLMs to map seizure semiology to potential EZ.

This study serves as an important reference for employing EpiSemoLLM in seizure semiology interpretation while highlighting its current constraints. With the ongoing development of LLMs and the availability of more training samples and importantly the inclusion of data from EEG, brain imaging and neuropsychology, the reliability and accuracy of EpiSemoLLM are expected to improve in the foreseeable future.

## Data Availability

All data produced in the present work are contained in the manuscript

## Acknowledgments

We acknowledge all epileptologists who participated in this study and all the insightful discussions.

## Notes

### Competing Interest Statement

The authors have declared no competing interest.

### Summary of Updates

The manucript is updated with modifications.

## References

1. Singh, A., Trevick, S.: The epidemiology of global epilepsy. Neurologic Clinics 34(4), 837–847 (2016)

2. Gavvala, J.R., Schuele, S.U.: New-onset seizure in adults and adolescents: a review. Jama 316(24), 2657–2668 (2016)

3. Chen, Z., Brodie, M.J., Liew, D., Kwan, P.: Treatment outcomes in patients with newly diagnosed epilepsy treated with established and new antiepileptic drugs: a 30-year longitudinal cohort study. JAMA neurology 75(3), 279–286 (2018)

4. Jan, M.M., Girvin, J.P.: Seizure semiology: value in identifying seizure origin. Canadian Journal of Neurological Sciences 35(1), 22–30 (2008)

5. Rosenow, F. and Lüders, H.: Presurgical evaluation of epilepsy. Brain, 124(9), pp.1683–1700 (2001)

6. Tufenkjian, K., Lüders, H.O.: Seizure semiology: its value and limitations in localizing the epileptogenic zone. Journal of clinical neurology 8(4), 243– 250 (2012)

7. Hirosawa, T., Kawamura, R., Harada, Y., Mizuta, K., Tokumasu, K., Kaji, Y., Suzuki, T., Shimizu, T.: ChatGPT-generated differential diagnosis lists for complex case–derived clinical vignettes: Diagnostic accuracy evaluation. JMIR Medical Informatics 11, 48808 (2023)

8. Wu, T., He, S., Liu, J., Sun, S., Liu, K., Han, Q.-L., Tang, Y.: A brief overview of ChatGPT: The history, status quo and potential future development. IEEE/CAA Journal of Automatica Sinica 10(5), 1122–1136 (2023)

9. Thirunavukarasu, A.J., Ting, D.S.J., Elangovan, K., Gutierrez, L., Tan, T.F., Ting, D.S.W.: Large language models in medicine. Nature medicine 29(8), 1930– 1940 (2023)

10. Zakka, C., Shad, R., Chaurasia, A., Dalal, A.R., Kim, J.L., Moor, M., Fong, R., Phillips, C., Alexander, K., Ashley, E., et al.: Almanac—retrieval-augmented language models for clinical medicine. NEJM AI 1(2), 2300068 (2024)

11. Benary, M., Wang, X.D., Schmidt, M., Soll, D., Hilfenhaus, G., Nassir, M., Sigler, C., Knoller, M., Keller, U., Beule, D., et al.: Leveraging large language models for decision support in personalized oncology. JAMA Network Open 6(11), 2343689– 2343689 (2023)

12. Liu, J., Wang, C., Liu, S.: Utility of ChatGPT in clinical practice. Journal of Medical Internet Research 25, 48568 (2023)

13. Johnson, D., Goodman, R., Patrinely, J., Stone, C., Zimmerman, E., Donald, R., Chang, S., Berkowitz, S., Finn, A., Jahangir, E., et al.: Assessing the accuracy and reliability of AI-generated medical responses: an evaluation of the chat-gpt model. Research square (2023)

14. Li, Y., Li, Z., Zhang, K., Dan, R., Jiang, S., Zhang, Y.: Chatdoctor: A medical chat model fine-tuned on a large language model meta-ai (llama) using medical domain knowledge. Cureus 15(6) (2023)

15. Xie, Q., Chen, Q., Chen, A., Peng, C., Hu, Y., Lin, F., Peng, X., Huang, J., Zhang, J., Keloth, V., et al.: Me llama: Foundation large language models for medical applications. arXiv preprint 2402.12749 (2024)

16. Labrak, Y., Bazoge, A., Morin, E., Gourraud, P.-A., Rouvier, M., Dufour, R.: Biomistral: A collection of open-source pretrained large language models for medical domains. arXiv preprint 2402.10373 (2024)

17. Jiang, A.Q., Sablayrolles, A., Mensch, A., Bamford, C., Chaplot, D.S., Casas, D.d.l., Bressand, F., Lengyel, G., Lample, G., Saulnier, L., et al.: Mistral 7B. arXiv preprint 2310.06825 (2023)

18. Canese, K., Weis, S.: Pubmed: the bibliographic database. The NCBI handbook 2(1) (2013)

19. Alim-Marvasti, A., Romagnoli, G., Dahele, K., Modarres, H., Pérez-García, F., Sparks, R., Ourselin, S., Clarkson, M.J., Chowdhury, F., Diehl, B., et al.: Probabilistic landscape of seizure semiology localizing values. Brain Communications 4(3), 130 (2022)

20. Computational Neuroimaging, L.: FreeSurferWiki. https://surfer.nmr.mgh.harvard.edu/fswiki Accessed 2024-05-20

21. Klein, A., Tourville, J.: 101 labeled brain images and a consistent human cortical labeling protocol. Frontiers in neuroscience 6, 33392 (2012)

22. Taori, R., Gulrajani, I., Zhang, T., Dubois, Y., Li, X., Guestrin, C., Liang, P., Hashimoto, T.B.: Stanford alpaca: An instruction-following llama model (2023)

23. Hu, E.J., Shen, Y., Wallis, P., Allen-Zhu, Z., Li, Y., Wang, S., Wang, L., Chen, W.: Lora: Low-rank adaptation of large language models. arXiv preprint 2106.09685 (2021)

24. Kojima, T., Gu, S.S., Reid, M., Matsuo, Y., Iwasawa, Y.: Large language models are zero-shot reasoners. Advances in neural information processing systems 35, 22199–22213 (2022)

25. Yang, Z., Gan, Z., Wang, J., Hu, X., Lu, Y., Liu, Z., Wang, L.: An empirical study of gpt-3 for few-shot knowledge-based vqa. In: Proceedings of the AAAI Conference on Artificial Intelligence, vol. 36, pp. 3081–3089 (2022)

26. Meng, J., Luo, Y., Fotedar, N., Karakis, I., Rao, V.-R., Asmar, M., et al: Is ChatGPT better than epileptologists at interpreting seizure semiology? medRxiv, doi: 10.1101/2024.04.13.24305773 (2024)

27. Kim, H.-W., Shin, D.-H., Kim, J., Lee, G.-H., Cho, J.W.: Assessing the performance of chatgpt’s responses to questions related to epilepsy: A cross-sectional study on natural language processing and medical information retrieval. Seizure: European Journal of Epilepsy 114, 1–8 (2024)

28. Wu, Y., Zhang, Z., Dong, X., Hong, S., Hu, Y., Liang, P., Li, L., Zou, B., Wu, X., Wang, D., et al.: Evaluating the performance of the language model ChatGPT in responding to common questions of people with epilepsy. Epilepsy & Behavior 151, 109645 (2024)

29. Wu, C., Zhang, X., Zhang, Y., Wang, Y., Xie, W.: Pmc-llama: Further finetuning llama on medical papers. arXiv preprint 2304.14454 (2023)

30. Kamble, K., Alshikh, W.: Palmyra-med: Instruction-based fine-tuning of LLMs enhancing medical domain performance (2023)

31. Ryvlin P, Barba C, Bartolomei F, et al. Grading system for assessing the confidence in the epileptogenic zone reported in published studies: A Delphi consensus study. Epilepsia. 2024;65(5):1346–1359. doi:10.1111/epi.17928

